# A Novel Dual-Outcome Risk Calculator for Trial of Labor After Cesarean

**DOI:** 10.64898/2026.03.18.26348725

**Authors:** Lauren Crabtree, Ciprian P. Gheorghe

**Affiliations:** Department of Gynecology and Obstetrics, Loma Linda University School of Medicine, Loma Linda, California

**Keywords:** TOLAC, VBAC, risk prediction model, maternal morbidity, neonatal morbidity

## Abstract

**Objective:** To develop and validate a multivariable prediction model and clinically actionable risk score for vaginal birth after cesarean (VBAC) success using machine learning, and to integrate neonatal morbidity outcomes into a decision-analytic framework for trial of labor after cesarean (TOLAC) counseling.

**Methods:** We performed a retrospective cohort study of 1,418 consecutive TOLAC cases at a single tertiary care center in California from 2019 through 2025. Multivariable logistic regression and four machine learning algorithms (logistic regression, random forest, gradient boosting, extreme gradient boosting) were trained using 5-fold stratified cross-validation. A cumulative risk score (negative 1 to 7 points) was constructed from independently significant predictors. Neonatal intensive care unit (NICU) admission rates and uterine rupture rates were evaluated across risk strata.

**Results:** The overall VBAC rate was 76.7% (1,087/1,418). Penalized logistic regression achieved the highest cross-validated AUC (0.71, 95% CI 0.67 to 0.75). A parsimonious multivariable logistic model used for score derivation had an AUC of 0.70 (95% CI 0.67 to 0.73). Independent predictors of failed TOLAC included induction of labor (adjusted odds ratio [aOR] 1.93, 95% CI 1.48 to 2.52), hypertensive disorders (aOR 1.60, 95% CI 1.19 to 2.15), diabetes mellitus (aOR 1.71, 95% CI 1.19 to 2.47), obesity (body mass index [BMI] 30 or greater; aOR 1.46, 95% CI 1.11 to 1.90), maternal age of 40 years or older (aOR 1.49, 95% CI 0.89 to 2.50), and gestational age of 41 weeks or greater (aOR 2.22, 95% CI 1.40 to 3.52). Prior vaginal delivery was independently protective (aOR 0.37, 95% CI 0.28 to 0.48). The cumulative risk score stratified VBAC success from 89.1% (score negative 1) to 37.8% (score 4 or higher). NICU admission rates increased concordantly from 31.7 to 200.0 per 1,000 across risk strata negative 1 through 4 or higher (Spearman rho 0.94, P for trend = .005). Uterine rupture occurred in 28 cases (1.97%) and was associated with severe maternal morbidity (10.7% vs 0.7%; odds ratio 16.56, P < .001) but was not predicted by any antepartum risk factor. Exclusion of patients with risk scores of 3 or higher (11.3% of the cohort) improved overall VBAC success to 80.0% (P = .04) and reduced NICU rates to 66.0 per 1,000.

**Conclusion:** A machine learning to derived cumulative risk score incorporating prior vaginal delivery as a protective factor identifies TOLAC candidates with poor VBAC prognosis and elevated neonatal morbidity, providing an evidence-based tool for individualized delivery counseling. Uterine rupture remains unpredictable by antepartum characteristics.

## INTRODUCTION

Trial of labor after cesarean (TOLAC) delivery is a crucial shared decision in obstetric practice. When successful, vaginal birth after cesarean (VBAC) reduces the risk of morbidity in future pregnancies. When unsuccessful, however, failed TOLAC confers risk that exceeds that of elective repeat cesarean.^1–3^ The challenge for clinicians is that the probability of success varies substantially across individuals, and no single predictor is sufficient to guide this decision.

Population-level VBAC success rates in the United States range from 60% to 80%, with considerable variation attributable to patient selection, institutional practice patterns, and the institutional support and policies.^1^ Several prediction models have been proposed, the most widely cited being the Grobman model derived from the Maternal-Fetal Medicine Units Network (MFMU), which incorporates maternal age, pre-pregnancy height and weight, treated chronic hypertension, indication for previous cesarean delivery, and obstetrical history.^4–6^ Although this model demonstrates reasonable discrimination, it was developed from data collected between 1999 and 2002, before the increases in maternal age and pre-pregnancy BMI that characterize the contemporary obstetric population.^4–6^ In the current era, when more patients enter pregnancy with advanced maternal age, obesity, and other comorbidities, there is a need to update and refine predictors of TOLAC success so that counseling accurately reflects the risk profile of today’s medically complex population.^4,7–8^ Despite its widespread use, the MFMU VBAC Calculator has notable limitations for contemporary counseling, as it reports broad probability ranges, which may constrain its precision and clinical applicability.

Machine learning methods offer potential advantages over traditional logistic regression for this classification task, as they can capture nonlinear interactions between maternal characteristics and labor management without requiring explicit specification of interaction terms. Recent applications of ensemble methods to obstetric prediction have shown promise.^10^ Extending these approaches to explicitly model how maternal characteristics and antepartum factors jointly shape TOLAC success probabilities may yield more clinically actionable, patient-specific counseling tools for an increasingly medically complex maternal population.

A further limitation of existing models is the singular focus on VBAC success as the outcome of interest. From the perspective of patient counseling, the relevant question is not merely whether vaginal delivery will be achieved, but whether the overall maternal and neonatal outcome profile favors TOLAC over elective repeat cesarean for a given patient. Neonatal morbidity represents a clinically meaningful endpoint that has not been incorporated into VBAC prediction frameworks or TOLAC decision aids. Incorporating neonatal outcomes alongside maternal risks is particularly important in contemporary obstetric populations with higher baseline risk profiles, where small differences in intrapartum management may substantially affect neonatal resource use and early-life morbidity.

We sought to integrate machine learning prediction and neonatal outcome data to develop a composite risk stratification tool that concurrently estimates the probability of VBAC success and the expected neonatal morbidity burden for individual TOLAC candidates. Further, we aimed to determine whether uterine rupture, the most catastrophic complication of TOLAC, can be predicted using antepartum characteristics and incorporated into a unified, clinically interpretable decision model to guide counseling in contemporary practice.

## MATERIALS AND METHODS

### Study Design and Population

We performed a retrospective cohort study of all patients who underwent TOLAC at a single tertiary care center in California from April 2019 through December 2025. The dataset was obtained from the Loma Linda University Children’s Hospital perinatal quality database. The original dataset included 1,419 consecutive cases with variables encompassing maternal demographics, clinical characteristics, labor parameters, delivery outcomes, and neonatal metrics. After excluding one case with a biologically implausible BMI (5,540 kg/m2), the final analytic cohort consisted of 1,418 TOLAC attempts. This study was approved by the institutional review board (IRB#5260029).

### Outcome Definitions

The primary outcome was successful VBAC, defined as vaginal delivery (spontaneous, vacuum-assisted, or forceps-assisted) after a prior cesarean. Secondary outcomes included neonatal intensive care unit (NICU) admission, severe maternal morbidity (SMM, excluding transfusion-only cases per Centers for Disease Control and Prevention criteria), and uterine rupture (identified by International Classification of Diseases, Tenth Revision [ICD-10] code O71.1, Rupture of uterus during labor, within the composite diagnosis field).

### Predictor Variables

Candidate predictors available at the time of delivery counseling included maternal age (years), pre-pregnancy BMI (kg/m2), gestational age (weeks), parity, race and ethnicity (self-reported), labor onset type (spontaneous vs induced), diabetes mellitus (none, gestational, pre-gestational), hypertensive disorders (none, chronic hypertension, preeclampsia-eclampsia), premature rupture of membranes (PROM), chorioamnionitis, and prior vaginal delivery (inferred from parity greater than 1, as patients with more than one prior cesarean are generally not offered TOLAC). Continuous variables were parsed from text fields using regular expression extraction and validated against source records.

### Statistical Analysis

Bivariate associations between candidate predictors and VBAC success were assessed using chi-square or Fisher exact tests for categorical variables and Mann-Whitney U tests for continuous variables with nonnormal distributions. Multivariable logistic regression was performed using backward elimination with a significance threshold of P < 0.10. Results were reported as adjusted odds ratios (aOR) with 95% confidence intervals (CI). Model calibration was evaluated using the Hosmer-Lemeshow goodness-of-fit test.

### Machine Learning

Four machine learning algorithms were evaluated: penalized logistic regression (L2 regularization), random forest (500 trees, maximum depth of 6), gradient boosting (200 estimators, learning rate of 0.1), and extreme gradient boosting (XGBoost). Each model was trained using 5-fold stratified cross-validation to mitigate overfitting. Hyperparameters were selected based on the cross-validated area under the receiver operating characteristic curve (AUC). Feature importance was derived from the random forest model using the mean decrease in Gini impurity, and model calibration was visualized by calibration plots.

### Cumulative Risk Score

A clinically applicable risk score was developed from independent predictors identified in the multivariate logistic regression and machine learning, weighted by the magnitude and direction of their associations. Points were assigned as follows: BMI of 30 or greater (1 point), BMI of 40 or greater (1 additional point), induction of labor (1 point), diabetes mellitus (1 point), hypertensive disorder (1 point), maternal age of 40 years or older (1 point), and gestational age of 41 weeks or greater (1 point). Prior vaginal delivery was assigned negative 1 point, yielding a total score range of negative 1 to 7. VBAC success rates and NICU admission rates were calculated at each score level with 95% Wilson confidence intervals (CI). The Spearman rank correlation coefficient was used to test for monotonic trend across grouped risk strata (scores negative 1, 0, 1, 2, 3, and 4 or higher).

### Exclusion Threshold Analysis

To evaluate the clinical utility of the risk score for identifying patients who should be counseled toward elective repeat cesarean delivery, we performed a sequential exclusion analysis comparing VBAC success rates, NICU admission rates (per 1,000 deliveries), and uterine rupture rates (per 1,000 deliveries) across three cohorts: all TOLAC patients, patients with risk score less than 3, and patients with risk score less than 2. Pairwise comparisons were conducted using two-proportion Z tests. The characteristics of excluded patients were analyzed separately to quantify the counseling trade-off.

All analyses were performed using Python version 3.10 with NumPy, pandas, SciPy, statsmodels, scikit-learn, and XGBoost. A two-tailed P value less than 0.05 was considered statistically significant. Random seeds were fixed at 42 to ensure reproducibility. Complete code is provided in the Supplemental Methods. Claude was used to assist with development of Python code for statistical analyses, and all resulting code was reviewed, verified, and modified as needed by the authors.

## RESULTS

### Study Population

The analytic cohort included 1,418 patients who attempted TOLAC. The median maternal age was 31 years (interquartile range [IQR] 27 to 35), median pre-pregnancy BMI was 28.4 kg/m2 (IQR 24.8 to 33.3), and median gestational age at delivery was 39.1 weeks (IQR 38.4 to 40.0). The cohort was predominantly Hispanic (63.1%; 43.8% U.S.-born, 19.3% non to U.S.-born), followed by White (15.8%), Black (8.7%), multiracial (5.3%), Asian (4.0%), and other (3.1%). Labor was induced in 615 cases (43.4%). Among study participants, 178 (12.6%) had diabetes mellitus, 342 (24.1%) had hypertensive disorders, 93 (6.6%) had PROM, and 104 (7.3%) developed chorioamnionitis. Prior vaginal delivery, inferred from parity greater than 1, was present in 826 patients (58.3%). Demographic and clinical characteristics are presented in Table 1.

**Table 1.** Demographic and clinical characteristics of the study population. (A) Body mass index category distribution. (B) Race and ethnicity. (C) Clinical factor prevalence by VBAC outcome. (D) Annual VBAC trend. (E) Birth weight distributions. (F) Body mass index distributions by delivery outcome.

| Characteristic | VBAC Success (n = 1,087) | Failed TOLAC (n = 331) | P Value |
| --- | --- | --- | --- |
| Maternal age, y, median (IQR) | 31.0 (27.0 to 35.0) | 31.0 (28.0 to 35.0) | .422 |
| BMI, kg/m<sup>2</sup>, median (IQR) | 28.1 (24.4 to 32.7) | 30.2 (26.0 to 35.3) | < .001 |
| Gestational age, wk, median (IQR) | 39.1 (38.4 to 40.0) | 39.3 (38.6 to 40.3) | .003 |
| Induction of labor, n (%) | 418 (38.5) | 197 (59.5) | < .001 |
| Diabetes mellitus, n (%) | 120 (11.0) | 58 (17.5) | .003 |
| Hypertensive disorder, n (%) | 227 (20.9) | 115 (34.7) | < .001 |
| Obesity (BMI ≥ 30), n (%) | 403 (37.1) | 166 (50.2) | < .001 |
| Morbid obesity (BMI ≥ 40), n (%) | 58 (5.3) | 34 (10.3) | .002 |
| Prior vaginal delivery, n (%) | 693 (63.8) | 133 (40.2) | < .001 |
| Maternal age ≥ 40 y, n (%) | 68 (6.3) | 25 (7.6) | .479 |
| GA ≥ 41 wk, n (%) | 60 (5.5) | 36 (10.9) | .001 |
| NICU admission, n (%) | 57 (5.2) | 46 (13.9) | < .001 |

Data are presented as median (interquartile range) or n (%). P values from Mann-Whitney U test for continuous variables and chi-square test for categorical variables.

### Primary Outcome: VBAC Success

Successful VBAC was achieved in 1,087 of 1,418 cases (76.7%), including 1,008 spontaneous vaginal deliveries and 79 operative vaginal deliveries (78 vacuum, 1 forceps). Repeat cesarean delivery was performed in 331 cases (23.3%). The annual VBAC rate remained stable across the study period (range 73.9% to 78.7%).

### Bivariate Predictors of VBAC

In bivariate analysis, VBAC success was associated with lower BMI (mean 28.9 vs 31.2 kg/m2, *p* < .001), spontaneous labor (83.3% vs 68.0% for induction, *p* < .001), absence of diabetes (78.0% vs 67.4%, *p* < .001), absence of hypertensive disorders (79.9% vs 66.4%, *p* < .001), and absence of chorioamnionitis (77.9% vs 60.6%, P *=* .02). Prior vaginal delivery was associated with higher VBAC success (83.9% vs 66.6%, P < .001). Race and ethnicity, maternal age, gestational age, and premature rupture of membranes were not significantly associated with VBAC success in bivariate analysis.

### Multivariable Logistic Regression

On multivariable analysis, seven predictors were retained in the final model. Risk factors for failed TOLAC included gestational age of 41 weeks or greater (aOR 2.22, 95% CI 1.40 to 3.52), induction of labor (aOR 1.93, 95% CI 1.48 to 2.52), diabetes mellitus (aOR 1.71, 95% CI 1.19 to 2.47), hypertensive disorders (aOR 1.60, 95% CI 1.19 to 2.15), maternal age of 40 years or older (aOR 1.49, 95% CI 0.89 to 2.50), and obesity (BMI 30 or greater; aOR 1.46, 95% CI 1.11 to 1.90). Prior vaginal delivery was independently protective (aOR 0.37, 95% CI 0.28 to 0.48). The model demonstrated adequate calibration (Hosmer-Lemeshow P *=* 0.79) and an overall AUC of 0.70 (95% CI 0.67 to 0.73; Figure 1).

**Figure 1.**
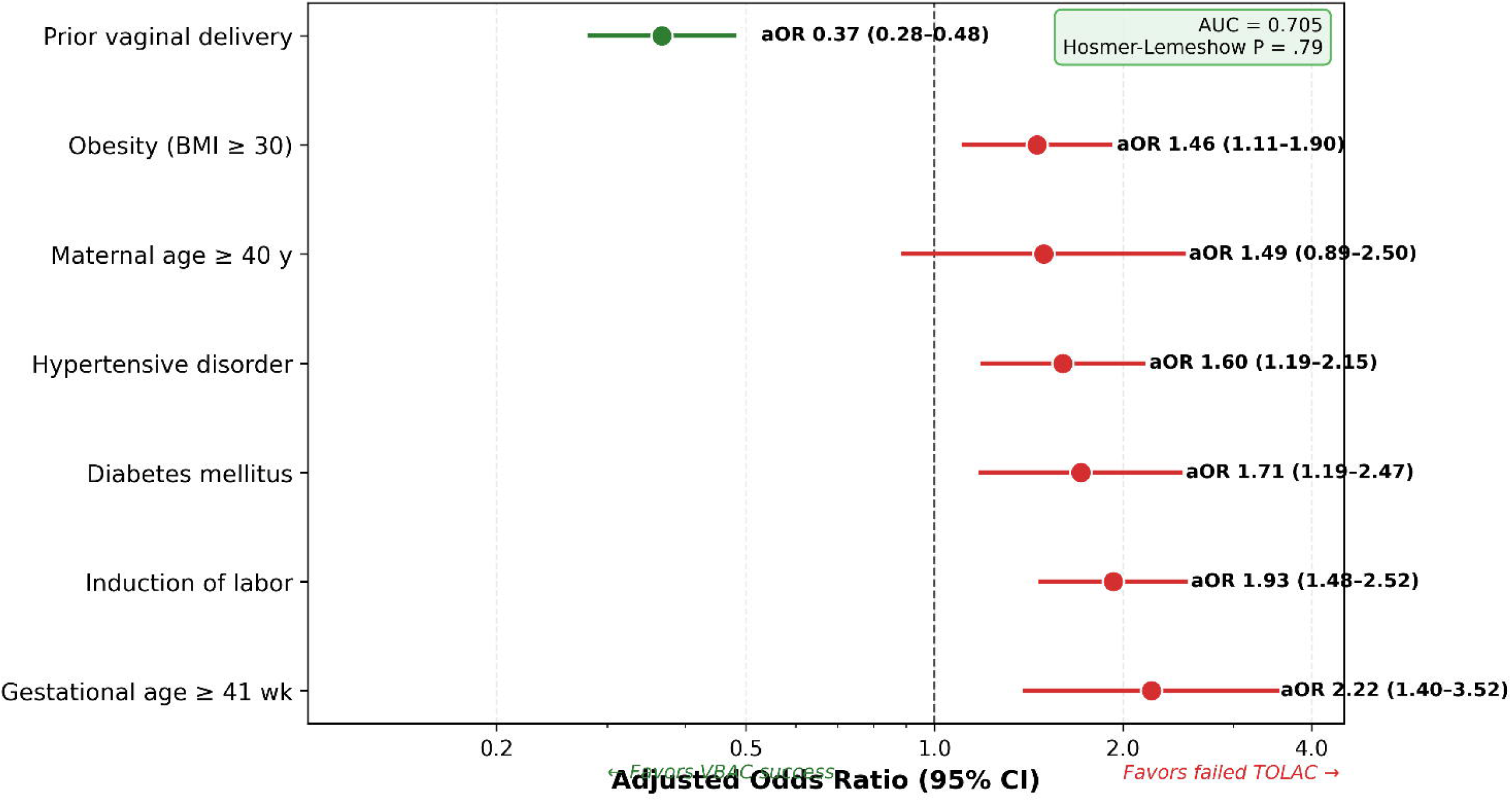
Forest plot of adjusted odds ratios from multivariable logistic regression for prediction of successful VBAC. Error bars represent 95% confidence intervals. The vertical dashed line indicates the null value (odds ratio 1.0).

### Machine Learning Performance

Among the four machine learning algorithms evaluated, penalized logistic regression achieved the highest cross-validated AUC (0.71; 95% CI 0.68 to 0.74), followed by random forest (0.70; 95% CI 0.66 to 0.73), gradient boosting (0.67; 95% CI 0.63 to 0.70), and XGBoost (0.67; 95% CI 0.63 to 0.70). Feature importance analysis from the random forest model identified BMI, prior vaginal delivery, gestational age, labor type, and maternal age as the strongest predictors of VBAC success. All models demonstrated acceptable calibration, with predicted probabilities closely aligning with observed event rates across deciles (Figure 2).

**Figure 2.**
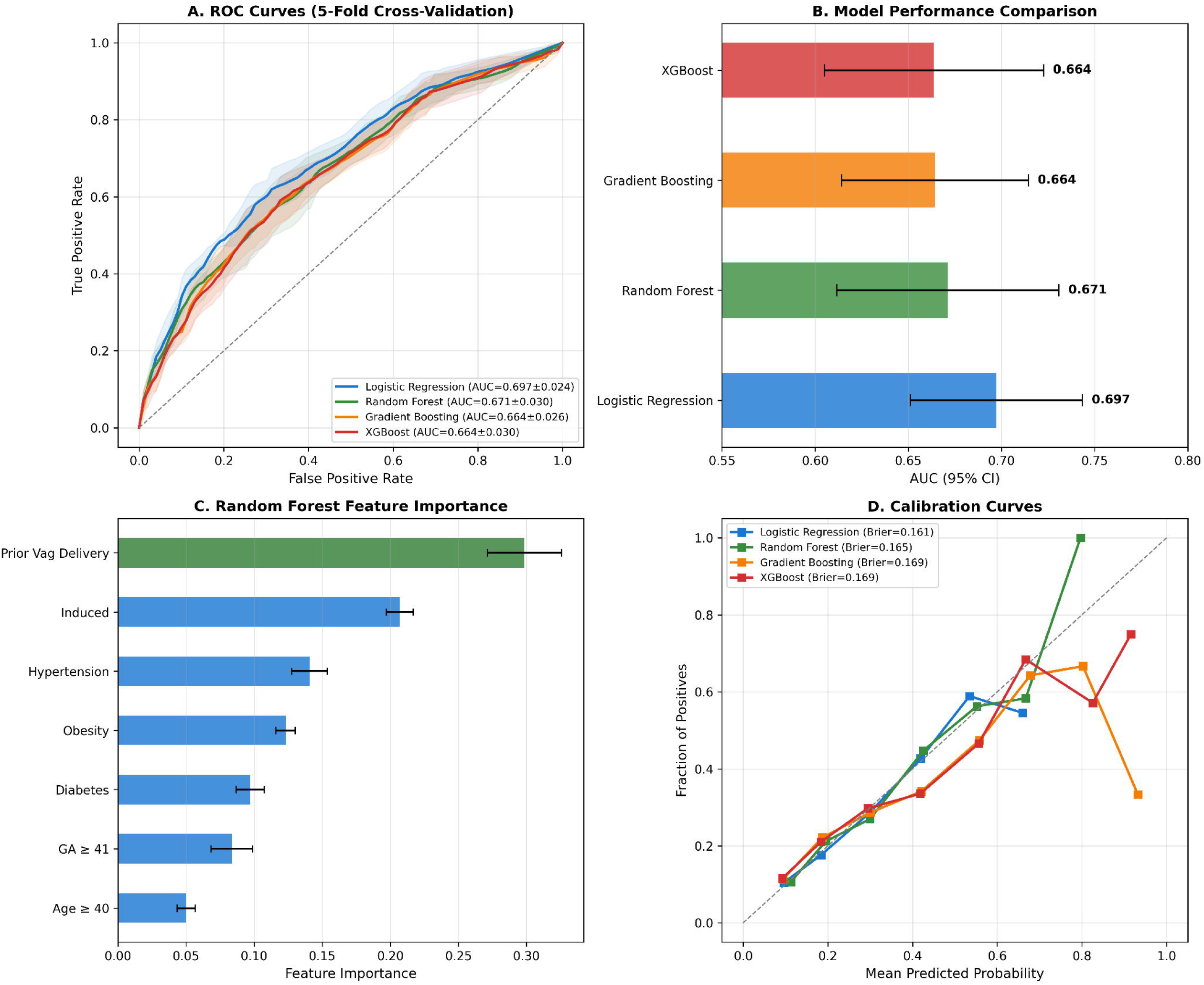
Machine learning analysis. (A) Receiver operating characteristic curves for all four models. (B) Cross-validated area under the curve comparison. (C) Random forest feature importance by mean decrease in Gini impurity. (D) Calibration curves by decile of predicted probability.

### Cumulative Risk Score

The cumulative risk score demonstrated a monotonic relationship with VBAC success across the negative 1 to 4+ range: score negative 1 (89.1%, n = 221), score 0 (86.0%, n = 420), score 1 (78.0%, n = 386), score 2 (63.6%, n = 231), score 3 (55.7%, n = 115), and score 4 or higher (37.8%, n = 45; Spearman rho negative 1.00, *p* < .001). Patients with scores of 3 or higher (n = 160, 11.3% of the cohort) had a combined VBAC rate of 50.6%, a threshold at which the risk-benefit balance of TOLAC becomes less favorable (Figure 3).

**Figure 3.**
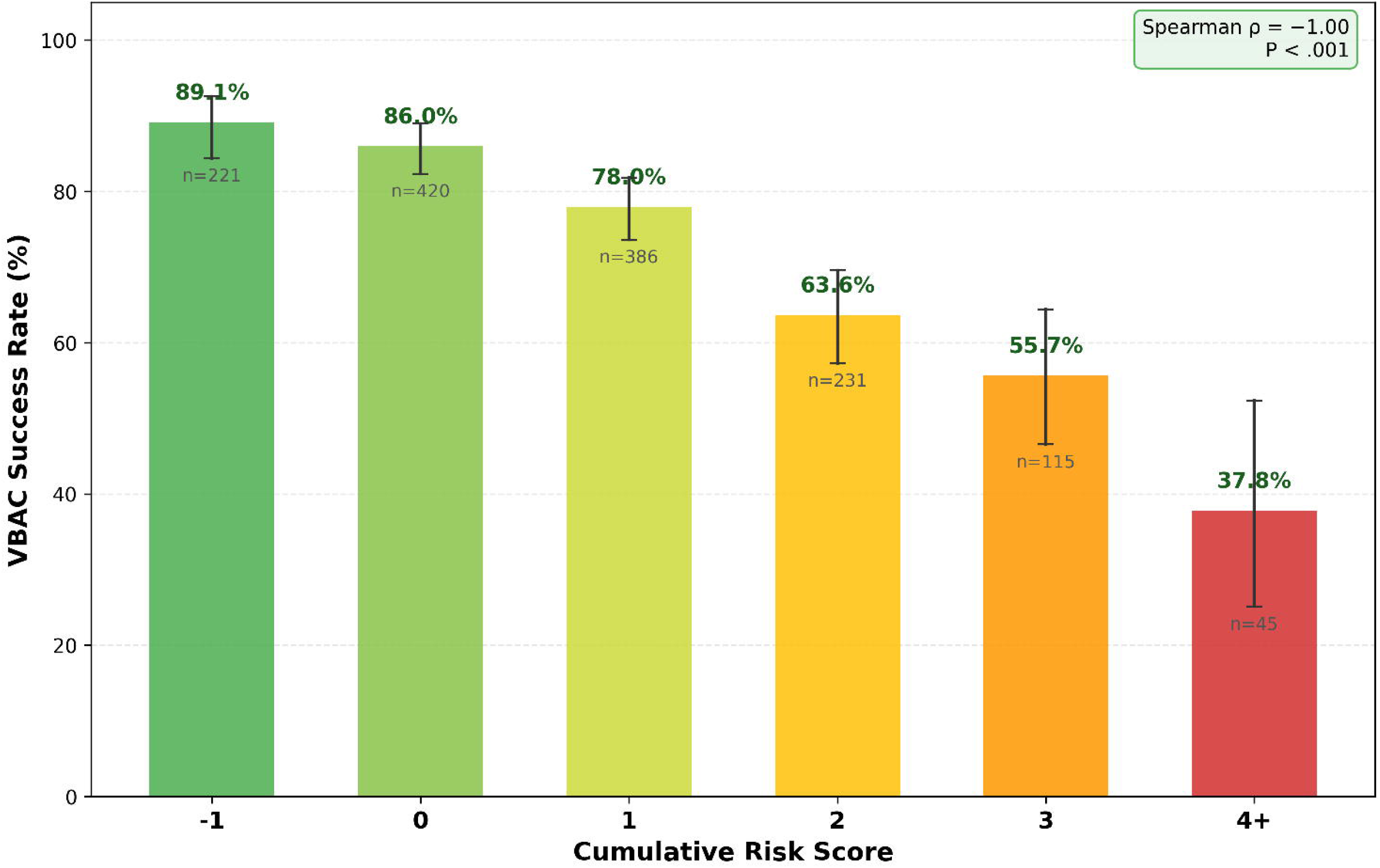
Risk stratification. (A) VBAC success rate by individual risk factor. (B) VBAC rate by number of concurrent risk factors. (C) VBAC rate by cumulative risk score (negative 1 to 4+) with 95% Wilson confidence intervals. (D) Body mass index by labor onset interaction matrix.

### Neonatal Outcomes by Risk Score

NICU admission rates increased concordantly with the cumulative risk score: 31.7 per 1,000 at score negative 1, 57.1 at score 0, 64.8 at score 1, 116.9 at score 2, 95.7 at score 3, and 200.0 at score 4 or higher (Spearman rho 0.94, *p* < .001). Scores of 5, 6, and 7 were observed in small numbers and were collapsed into the 4 or higher stratum for trend analysis. On multivariable analysis, independent predictors of NICU admission included chorioamnionitis (aOR 6.93, 95% CI 4.16 to 11.54), failed TOLAC (aOR 2.53, 95% CI 1.60 to 4.01), and operative vaginal delivery (aOR 2.42, 95% CI 1.09 to 5.36). The random forest machine learning model achieved the highest discrimination for NICU prediction (AUC 0.71, 95% CI 0.61 to 0.80; Figure 4).

**Figure 4.**
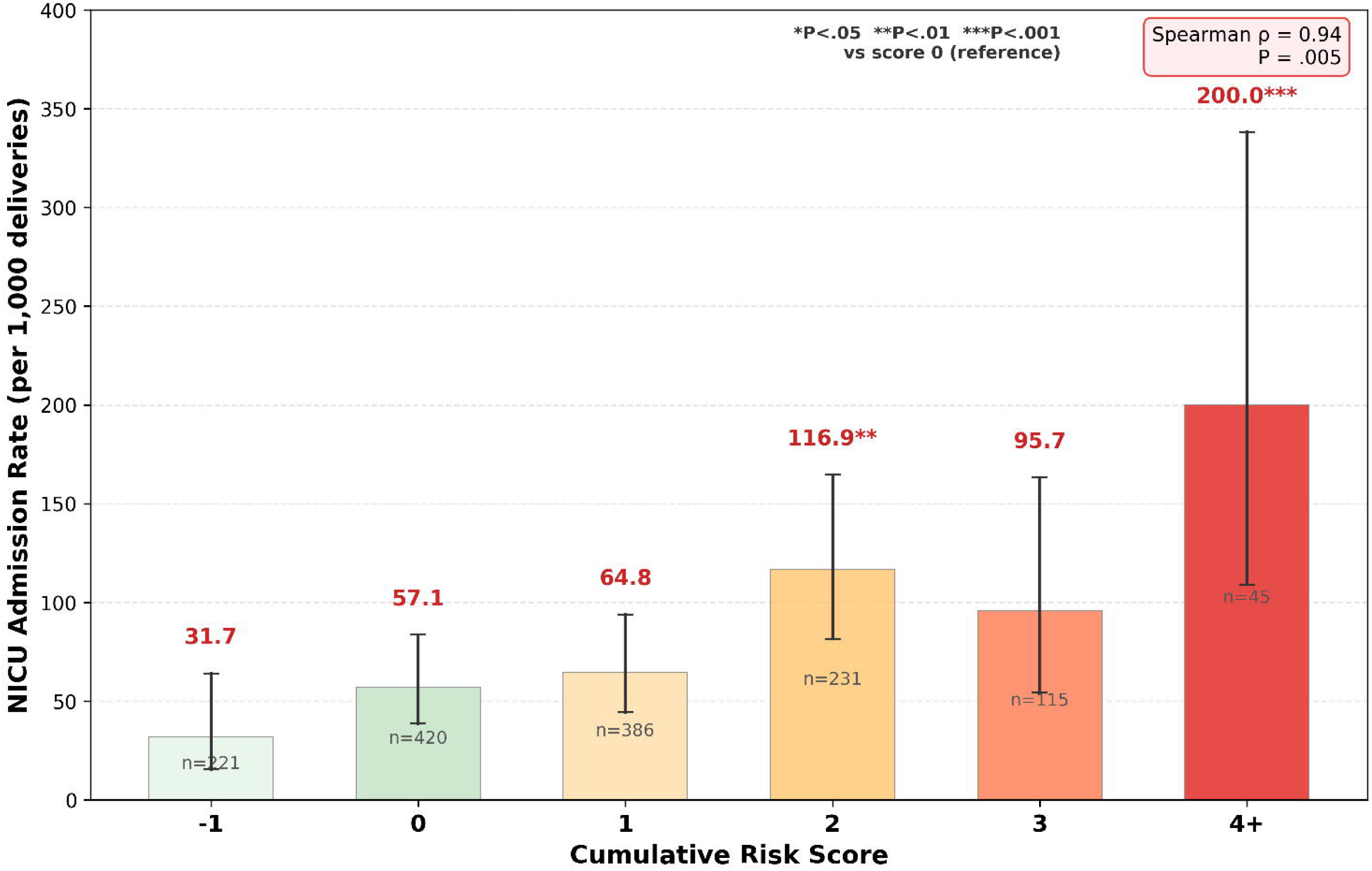
Neonatal intensive care unit admission analysis. (A) Forest plot of adjusted odds ratios. (B) Receiver operating characteristic curves. (C) Feature importance. (D) Model performance comparison.

### Uterine Rupture

Uterine rupture was identified in 28 cases (1.97%, 95% CI 1.37 to 2.84%). Among rupture cases, 25 (89.3%) underwent repeat cesarean delivery, and 3 (10.7%) achieved vaginal delivery. Severe maternal morbidity occurred in 3 of 28 rupture cases (10.7%) compared with 10 of 1,390 non-rupture cases (0.7%; odds ratio 16.56, 95% CI 4.32 to 63.42, P < .001). No maternal or antepartum characteristic was significantly associated with uterine rupture (all P > 0.10; Table 2).

**Table 2.**
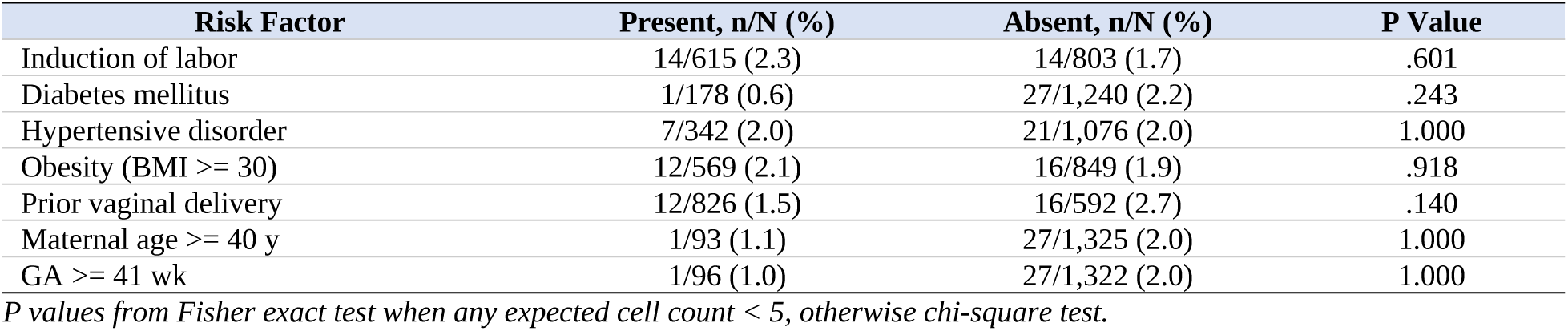
Uterine Rupture Rate by Risk Factor. Uterine rupture was identified in 28 of 1,418 cases (1.97%). No antepartum risk factor was significantly associated with rupture (all P > .10).

### Exclusion Threshold Analysis

Exclusion of patients with risk scores of 3 or higher (n = 160, 11.3% of the cohort) improved overall VBAC success from 76.7% to 80.0% (P *=* .04) and reduced NICU rates from 72.6 to 66.0 per 1,000 (P = .50). The more stringent exclusion of scores 2 or higher (n = 391, 27.6%) improved VBAC success to 83.6% (P < .001) and reduced NICU rates to 54.5 per 1,000 (*p* = .07). Uterine rupture rates were not significantly altered by either threshold (P = .94 and P = .87, respectively). Among the 160 excluded patients (score 3 or higher), the observed VBAC rate was 50.6% and the NICU rate was 125.0 per 1,000, both significantly different from the retained cohort (P < .001 and P = .007, respectively; Figure 5).

**Figure 5.**
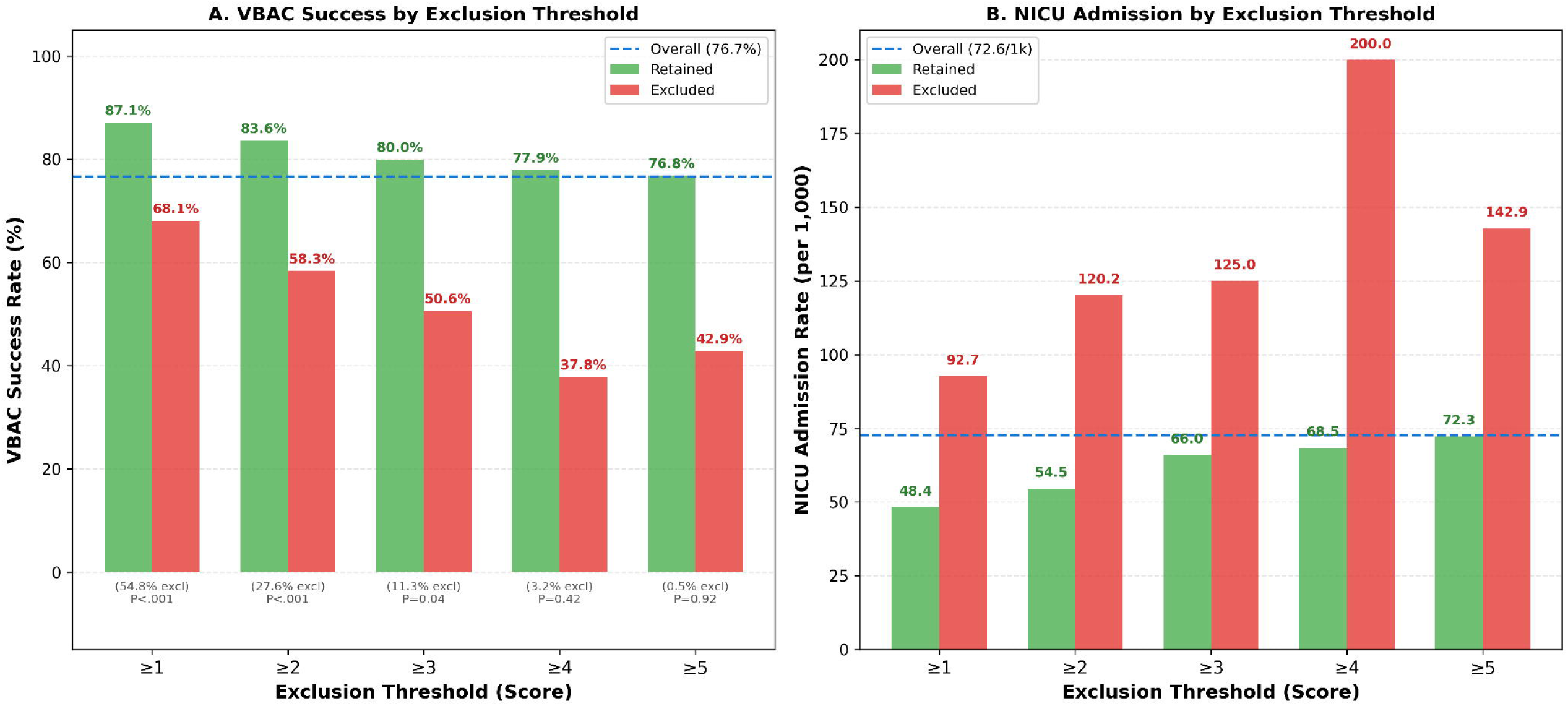
Exclusion threshold analysis. (A) VBAC success rate by exclusion threshold with pairwise P values. (B) NICU admission rate per 1,000 by exclusion threshold. (C) Uterine rupture rate per 1,000 by exclusion threshold. (D) Trade-off analysis showing number of patients excluded and their observed VBAC rate.

## COMMENT

Our study of VBAC success and neonatal outcomes at a single-center cohort has shown that a simple antepartum cumulative risk score effectively stratified both VBAC success and neonatal outcomes. By summing seven routinely available parameters into a negative 1 to 7 point score, we observed a graded decline in VBAC probability, with a parallel, clinically important rise in NICU admission rates. Thus, patients at higher scores are not only less likely to achieve VBAC; they also face greater neonatal morbidity. This dual gradient moves beyond the traditional binary focus on mode of delivery and aligns more closely with how patients conceptualize this decision, balancing their desire to avoid another cesarean against realistic expectations for their infant’s early course.

Our findings extend prior VBAC prediction work in several notable ways. Existing models, including the widely used MFMU calculator, were derived from earlier cohorts with lower prevalence of obesity and medical comorbidity, rely on more complex variable sets, and primarily estimate the chance of vaginal delivery without explicitly quantifying neonatal risk.^4–6^ In contrast, we created a tool for easy calculation and patient interpretation by restricting our score to comorbidities, obstetric history, and management decisions available at the time of counseling and linked each score level to both VBAC probability and NICU admission rates (Figure 6). Of note, prior vaginal delivery emerged as the strongest protective factor in both regression and machine learning analyses. Patients with a prior vaginal delivery had substantially higher VBAC success (83.9% vs 66.6%), and the inclusion of this variable as a negative 1 point in the score improved model discrimination and sharpened the identification of high-risk candidates. The pattern we observed, where higher risk scores concentrate both failed TOLAC and neonatal morbidity, supports using this cumulative risk measure not as a gatekeeper, but as a structured way to identify patients for whom elective repeat cesarean should be actively considered rather than reflexively dismissed. This score should allow for individualized counseling aligned with patient preferences and risk tolerance.

**Figure 6.**
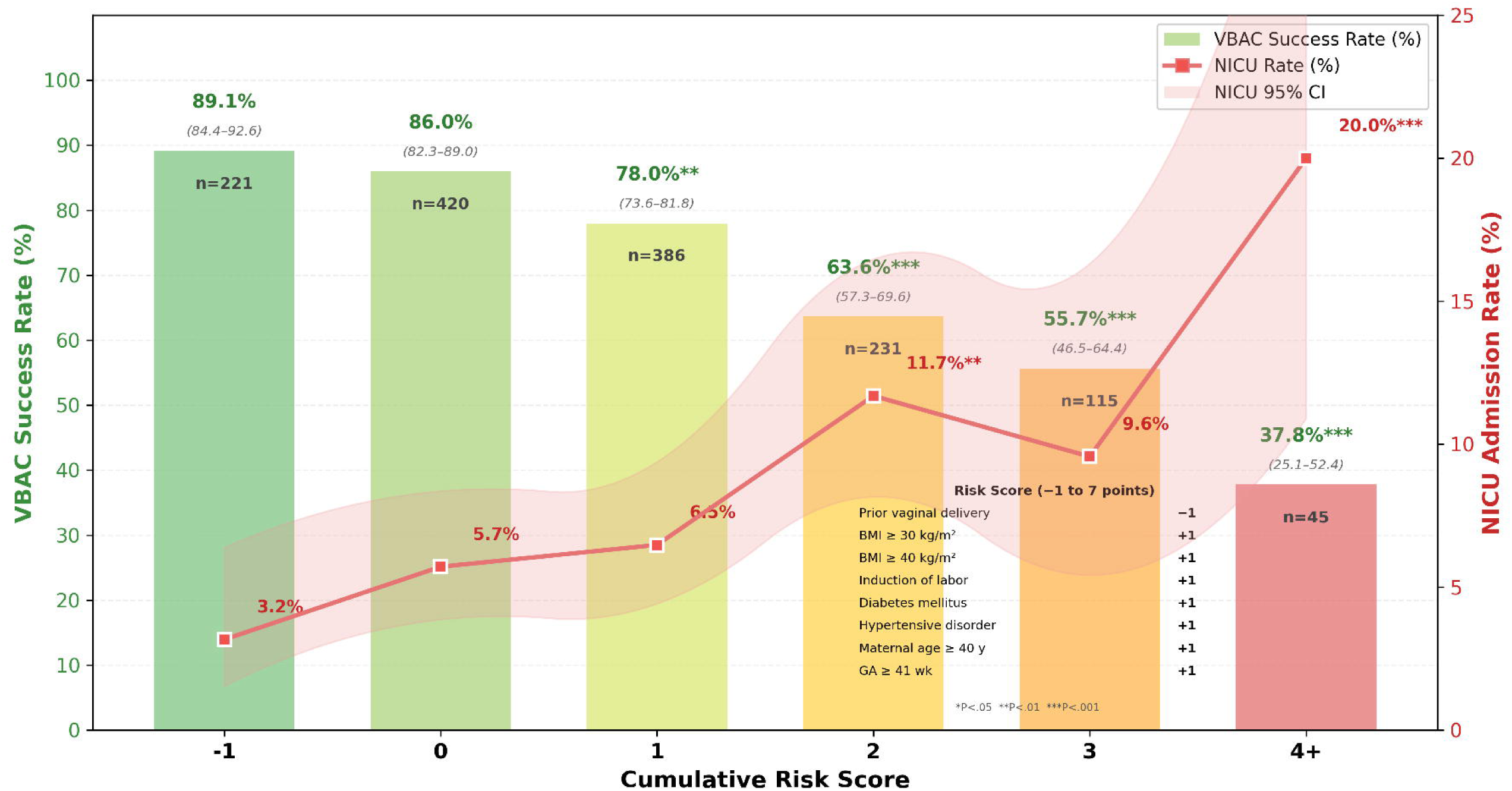
Dual-outcome risk score: VBAC success rate (%) and NICU admission rate (per 1,000) by cumulative risk score (negative 1 to 4+) with 95% Wilson confidence intervals.

An important counterpoint to the apparent precision of these models is our finding that despite extensive modeling, uterine rupture risk remained unpredictable and occurred more frequently than in some published cohorts.^12–13^ No individual variable or component of the cumulative score identified a subgroup at meaningfully lower or higher rupture risk in contrast to prior population-based work.^12–15^ Thus, counseling must continue to emphasize that rupture is an inherent risk of TOLAC, reinforcing the need for immediate surgical capability in all settings offering this option.

Derived from a contemporary, comorbidity-heavy cohort that resembles current TOLAC candidates, this study translates complex machine learning and decision-analytic work into a simple antepartum score based entirely on routinely available variables, and directly links each score level to both VBAC probability and NICU admission rates, enabling real-time, dual-outcome counseling at the bedside. However, these findings are limited by their derivation from practice patterns at a single tertiary center, the absence of key historical details such as indication and number of prior cesareans, and the small number of rare adverse events. These factors may limit the generalizability of the findings and underscore the need for external validation in diverse settings with varying TOLAC policies and NICU utilization. Future research should prospectively validate and integrate this cumulative risk score into decision-making tools, with the goal of shifting TOLAC counseling from a narrow focus on delivery mode to a more comprehensive, patient-centered dialog regarding maternal and neonatal outcomes that aligns delivery planning with the individual patient’s values and priorities.

## Data Availability

All data produced in the present study are available upon reasonable request to the authors

